# Transfer transcriptomic signatures for infectious diseases

**DOI:** 10.1101/2020.09.28.20203406

**Authors:** Julia di Iulio, Istvan Bartha, Roberto Spreafico, Herbert W. Virgin, Amalio Telenti

## Abstract

The modulation of the transcriptome is among the earliest responses to infection, and vaccination. However, defining transcriptome signatures of disease is challenging because logistic, technical and cost factors limit the size and representativeness of samples in clinical studies. These limitations lead to poor performance of signatures when applied to new datasets or varying study settings. Using a novel approach, we leverage existing transcriptomic signatures as classifiers in unseen datasets from prospective studies, with the goal of predicting individual outcomes. Machine learning allowed the identification of sets of genes, which we name transfer transcriptomic signatures, that are predictive across diverse datasets and/or species (rhesus to humans) and that are also suggestive of activated pathways and cell type composition. We demonstrate the usefulness of transfer signatures in two use cases: progression of latent to active tuberculosis, and severity of COVID-19 and influenza A H1N1 infection. The broad significance of our work lies in the concept that a small set of archetypal human immunophenotypes, captured by transfer signatures, can explain a larger set of responses to diverse diseases.

## Introduction

Infection and vaccination trigger a robust transcriptome response in tissues or in blood. These perturbations occur in the setting of a pre-existing immunophenotype in each individual characterized by a transcriptome that is regulated by genetics, the environment, the microbiome and virome, and prior infections (*1-7*). In the case of acute infections such as viral respiratory diseases, viral disease transmitted by arthropod vectors, or in chronic viral infections such as HIV, responses in peripheral blood mononucleolar cells (PBMC) may lead to the transcriptional deregulation of thousands of genes that vary significantly between individuals based on the status of their immune system at the time of infection (*8-14*). Cellular, biological and functional factors contribute to this aggregate transcriptomic response. For example, changes in cell composition, the nature of inflammatory responses, bystander effects of tissue damage and therapeutic intervention generate complex patterns of expression. The diversity of approaches used to investigate transcriptome responses – study design, timing, technical platform – also contribute to the patterns of expression across studies. Additional sources of noise in transcriptome profiles may result from the relatively small sample size that characterize many publications and the pervasive impact of batch effects (*15*). Given these variations, it is perhaps not surprising that consensus transcriptomic signatures that reliably operate across studies have been challenging to identify. This is important for designing prospective analyses of the human immunophenotype and to taking advantage of the wealth of legacy data from earlier work and data repositories, for example to establish host-response-based diagnostics (*9, 16*), with an emphasis on meta-analytical approaches (*17, 18*).

Because of the above considerations, there is significant interest in developing methods that reproducibly identify transcriptome profiles as biomarkers of disease susceptibility or prediction of vaccine responses. A key challenge is the generation of appropriate datasets for each pathogen and study endpoint. This effort requires considerable planning and resourcing. Once biomarkers are identified, they still need to undergo extensive validation in additional cohorts and applied in settings other than the population in which the study was originally conducted. Ideally, biomarkers would be so robust that they could be transferred across studies, and possibly across pathogens and species. For example, the severity of responses to viral respiratory infections could be predicted using a set of shared responses based on a strong deregulation of interferon-stimulated genes observed in the setting of different viral infections (*19*). Similarly, vaccine protection could be predicted using shared markers of response (*12*). Underlying these questions is the possibility of baseline human immunophenotypes that can be predictive of differential responses to various perturbations (*20, 21*). The overarching concept tested herein is that while the broad field of biomarkers - and specifically transcriptomics-based biomarkers - emphasizes specificity (to pathogen, perturbation and/or study endpoint), we hypothesize that there are sets of common responses having the desirable properties of generalization and transferability.

Here we identify patterns of gene response comprising transfer signatures that can be learned from deposited datasets and tested for predictive power in independent transcriptomes associated with clinical metadata. This work evaluates the performance of such signatures across pathogens and studies, including the validity of signatures learned from animal models for studies of human disease. We present two use cases of transfer signatures for infection and vaccination. Our work establishes the validity of this approach and explores the nature of human immunophenotypes. If generally applicable in additional studies, the methods described here may lead to rapid evolution of clinically and biologically relevant concepts in immunity and pathogenesis.

## Results

### Selection of signatures and datasets

We conceived this study under the assumption that the literature provides short lists of genes (here described as *“signatures”*) that are predictive of study outcomes across different biological settings. The study design is shown in **Fig. 1**. Published signatures – referred as “*literature signatures*” – may or may not be associated with full access to raw data/sequencing counts. Thus, we collected 148 literature signatures to support exploratory analysis, and raw data from 6 transcriptome studies to train and 3 transcriptome studies to test machine learning models (**Methods, Data S1**). The pairing of literature signatures, training and test datasets are depicted in **Fig. S1**. Our study focused on infectious diseases; thus, the literature signatures were obtained from papers investigating (i) infection with dengue, SARS-CoV-1, SARS-CoV-2, MERS-CoV, influenza A virus (IAV) H1N1, H5N1, H3N2, measles and respiratory syncytial virus (RSV) (referred as ‘*infection signatures*’, N=43), (ii) vaccine response to hepatitis B virus (HBV), IAV H1N1, H3N2 and/or influenza B virus (IBV) and against tuberculosis (TB) and simian immunodeficiency virus (SIV), (referred as ‘*vaccine signatures*’, N=13), (iii) progression to active TB (referred as ‘*TB signatures*’, N=20); (iv) we also explored the information that was encoded in collections of signatures that characterize cell composition (referred as ‘*cell type signatures*’, N=22) (*22*) and (v) biological states assessed through hallmark gene sets of the Molecular Signatures Database (referred as ‘*hallmark signatures*’, MSigDB, https://www.gsea-msigdb.org/gsea/msigdb/collections.jsp, N=50). The reason for including the latter two groups was to evaluate whether these signatures could be informative of the processes under study: cell type composition in bulk measuremnets is critical for the final aggregate readout; hallmark gene sets, because it is a compact representation of main biological processes/pathways. Of note, signatures were not restricted to human studies, as some were sourced from studies in rhesus and cynomolgus macaques (**Suppl. Data S1**).

**Fig. 1.**
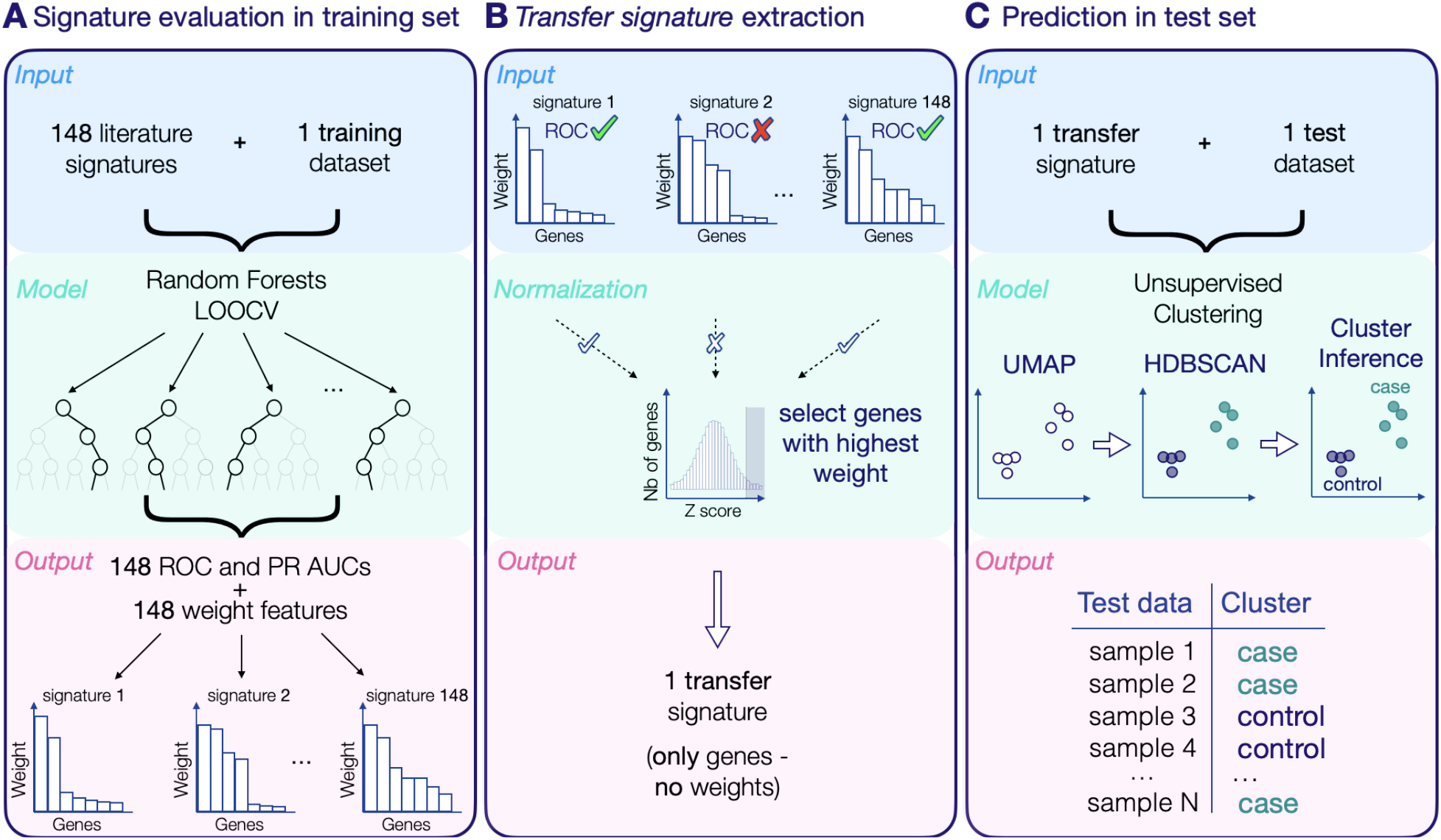
Study design. Three steps to progress from literature signatures to transfer signatures to prediction in unseen datasets. UMAP, Uniform Manifold Approximation and Projection. HDBSCAN, Hierarchical Density-Based Spatial Clustering of Application with Noise. LOOCV, leave-one-out cross validation. For additional information, see **Suppl. Fig. S1, Methods** and **Suppl. Data S1**.

Six publications provided different training datasets (**Suppl. Fig. S1**): one study on dengue infection (*23*), one study on IAV H1N1 infection (*24*), one study on trivalent influenza vaccination (comprising two cohorts, one with males and one with females (*25*)), one study on HBV vaccination (*26*) and one study on TB vaccination in rhesus macaques (*27*). Their descriptions and sources are provided in **Suppl. Data S1**. Of note, these studies contain multiple (N=14) non-independent datasets (timepoints). This design is expected to help understand the biology of transcriptome signatures and to monitor what are the earliest time points with predictive power.

### Training and testing of signatures

The study design uses random forest models to evaluate the collection of signatures on each training transcriptome datasets (**Fig. 1A, Suppl. Fig. S1, Suppl. Data S1**), followed by the extraction of a common set of predictive genes (*transfer signature*) from each training dataset (**Fig. 1B, Suppl. Fig. S1, Suppl. Data S1**) and finally using the transfer signature obtained from one training dataset to predict the outcome in an unseen, unrelated test datasets using unsupervised methods to exclude overfitting (**Fig. 1C, Suppl. Fig. S1, Suppl. Data S1**).

In the first step, we characterized the performance of all 148 literature signatures on each training dataset. For each dataset, we trained and evaluated machine learning models with the feature set restricted to the genes contained in each signature. Effectively, for each of the 14 training datasets (for example, dengue infection) we obtained 148 models, yielding a total of 2,072 models across the training data sets. Then, for each model, we computed the receiver operating characteristic (ROC) values and the individual importance of each gene. We computed the ROC area under the curve (AUROC) using the leave-one-out cross validation (LOOCV) strategy.

Because the percent split between cases and controls is different in each dataset, AUROCs could not be compared directly: for this reason, we expressed results as percentiles rather than raw AUROC. Percentiles were obtained by comparing the performance of the literature signatures to random lists of genes of identical size. We observed that a large proportion of literature signatures performed well across training datasets, supporting the notion that published signatures contain valuable shared information that can be used to train predictive models and classifiers (**Fig. 2A, Suppl. Table S1**). We compared the performance of the *cognate* signature (i.e., the signature that was specifically developed in the original study of the training dataset) for each training dataset against *literature* signatures (i.e., any other published signature considered in this study). All but one training dataset (HBV pre-vaccine) had at least one non-cognate signature outperforming the cognate signature in terms of ROC or precision-recall (PR) AUCs (**Suppl. Fig. S2**). These results supported the concept that the approach may identify signatures that can be transferred between datasets while retaining predictive power.

**Fig. 2.**
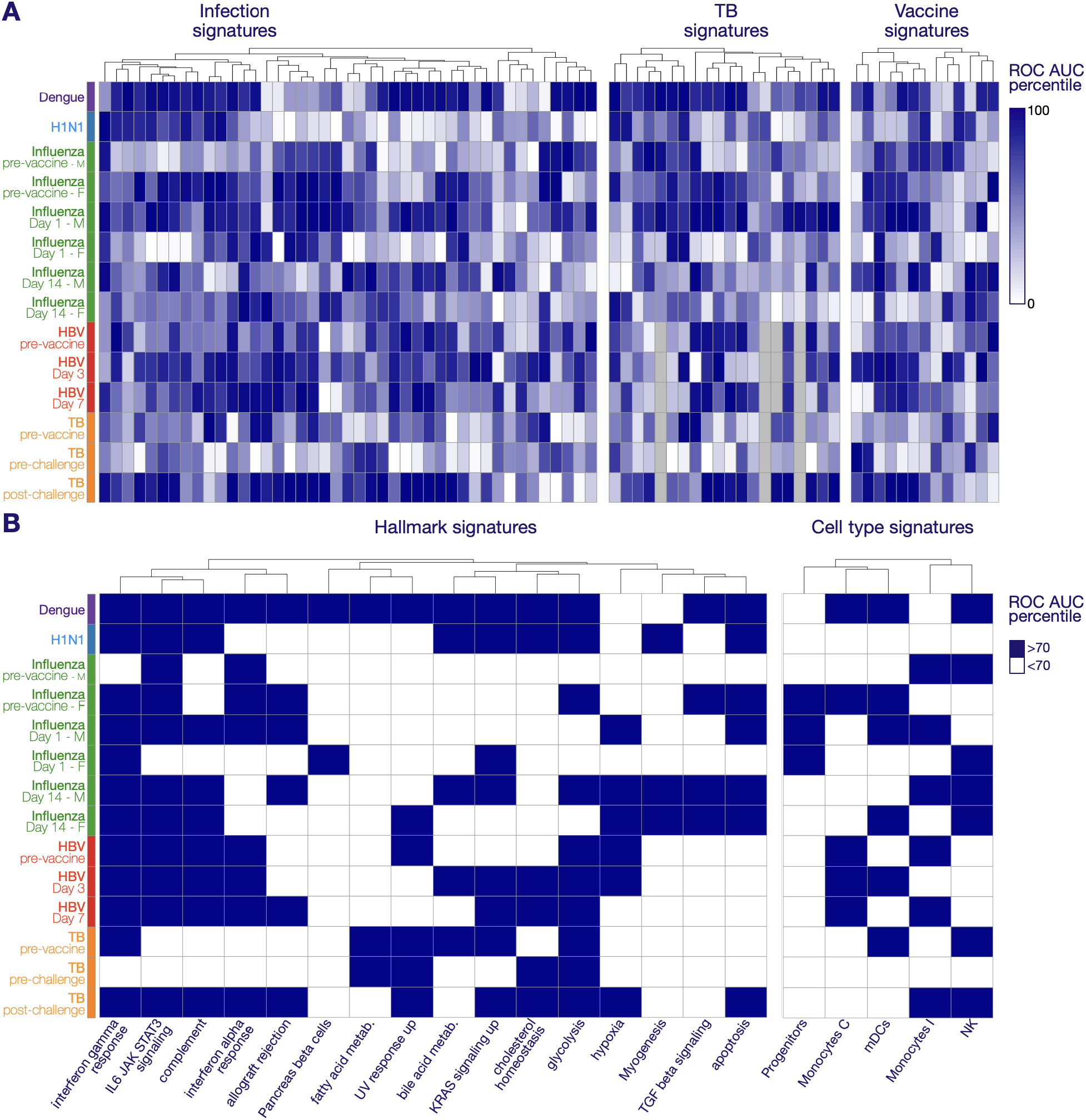
Performance of literature signatures. **Panel A** depicts a heatmap of the AUROCs obtained through radom forest models. Each column represents a signature from the literature, grouped by signature group. Each row represents a training dataset. In order to be able to compare the AUROC across the datasets (which do not have the same case/control distribution), the AUROC are depicted in percentiles. The percentiles are obtained by comparing the performance of the literature signature to 100 random gene lists of the same size. The higher the percentile (darker blue) the better the performance of the literature signature. Missing data is depicted in grey. The color annotation next to the rows indicates the infectious agent datasets. Influenza refers here to a tri-valent vaccine consisting of H1N1, H3N2 and IBV. **Panel B** displays the best performing hallmark and cell type signatures. Each row represents a training dataset (in the same order as in panel A). Columns represent the signatures - hallmark (left subpanel) and cell type (right panel) - that reached the 70^th^ percentile in at least one training dataset. For visual simplicity, the coloring here is binary as depicted in the legend. ROC, Receiver operating characteristic. AUC, area under the curve. Metab., metabolism. IL, interleukin. JAK, Janus kinase. STAT, signal transducer and activator of transcription. mDC, myeloid dendritic cell. NK, natural killer cell. F, female. M, male. For additional information, see **Methods** and **Suppl. Data S1**.

We also assessed two general sets of signatures, one sourced from Broad Institute’s MSigDB (“hallmark”) and one on cell type signatures (*22*), that are not explicitly designed for association with infection diseases (**Fig. 2A**). Despite that, a number of these general-purpose signatures also performed well with several datasets (**Suppl. Table S1**). As there is clear interest in understanding the nature of the signature that endows the generalization from hallmark and cell type signatures to other data sets, we further examined the signatures that performed at the top percentile in at least one dataset. **Fig. 2B** presents those top performing signatures across the various training datasets.

As several training datasets were time course experiments, we further inquired whether there existed patterns of performance across the hallmark and cell type signatures that would be consistent with current understanding of biology. We noticed that the relevance of particular signatures shifted according to the timepoint in the experiment. For example, in the rhesus macaque TB vaccine experiment (**Suppl. Data S1**, study 6), predictors of vaccine efficacy at baseline included hallmark signatures of interferon gamma response, fatty acid metabolism, UV response, bile acid metabolism, KRAS signaling and glycolysis, as well as cell type signatures of mDC and NK cells (**Fig. 2B**, TB pre-vaccine). In contrast, at the time of disease (TB post-challenge), the predictive signatures expanded to include hallmark IL6, JAK, STAT3 signaling, complement, interferon alpha response, allograft rejection, hypoxia and apoptosis, as well as a cell type signature of monocytes (**Fig. 2B**, TB post-challenge). The preeminence of these signatures is broadly consistent with current understanding of biomarkers, pathogenesis and cellular roles in tuberculosis(*28-32*). As another example, we observed differences in predictive signatures across gender category in the study of Franco et al.(*25*) that evaluated the response to influenza vaccination (**Fig. 2**). This is consistent with sex differences in the blood transcriptome associated with immune responses (*33, 34*).

Overall, the analysis of literature signatures from various sources support the original hypothesis that there are shared response elements that serve as biomarkers across multiple conditions. On this basis, we next sought to create signatures with predictive power across a wide range of diseases (‘transfer signatures’).

### Definition of transfer signatures

To establish a transfer signature for each training dataset, we used every literature signature that had a AUROC higher (heuristics) than the 70^th^ percentile compared to random lists of genes of identical size (**Methods, Fig. 1B**). This step creates 14 transfer signatures – one per training dataset. Importantly, for the purpose of defining transfer signatures, we excluded the cognate signature from this step in order to focus on genes that were also relevant in other published study. We then standardized the gene importance output from the random forest models of the signatures that passed the threshold and selected the first 50 genes (we also evaluated 10 and 20-gene signatures, **Suppl. Fig. S3**) with the highest standardized importance feature score (**Methods, Fig. 1B**). As expected – given that transfer signatures are made of the top-classifying genes for a given training dataset – transfer signatures performed well on the datasets they were trained on: AUROC varied between 0.85 and 0.97 and PR AUC of 0.72 to 0.98 for the various training datasets (**Fig. 3A** and **B)**. In all but one training dataset (TB pre-vaccine), they matched or improved the performance, in terms of AUROC, of the best single performing literature signature, including the cognate signature (**Suppl. Fig. S4**). The nature of the genes retained in the transfer signatures (listed in **Suppl. Data S1**) was assessed by network analysis. The resulting network had more interactions that expected (protein-protein enrichment p-value<1.0e^-16^); main functional enrichments prominently include immune system and metabolic processes (**Suppl. Fig. S5**).

**Fig. 3.**
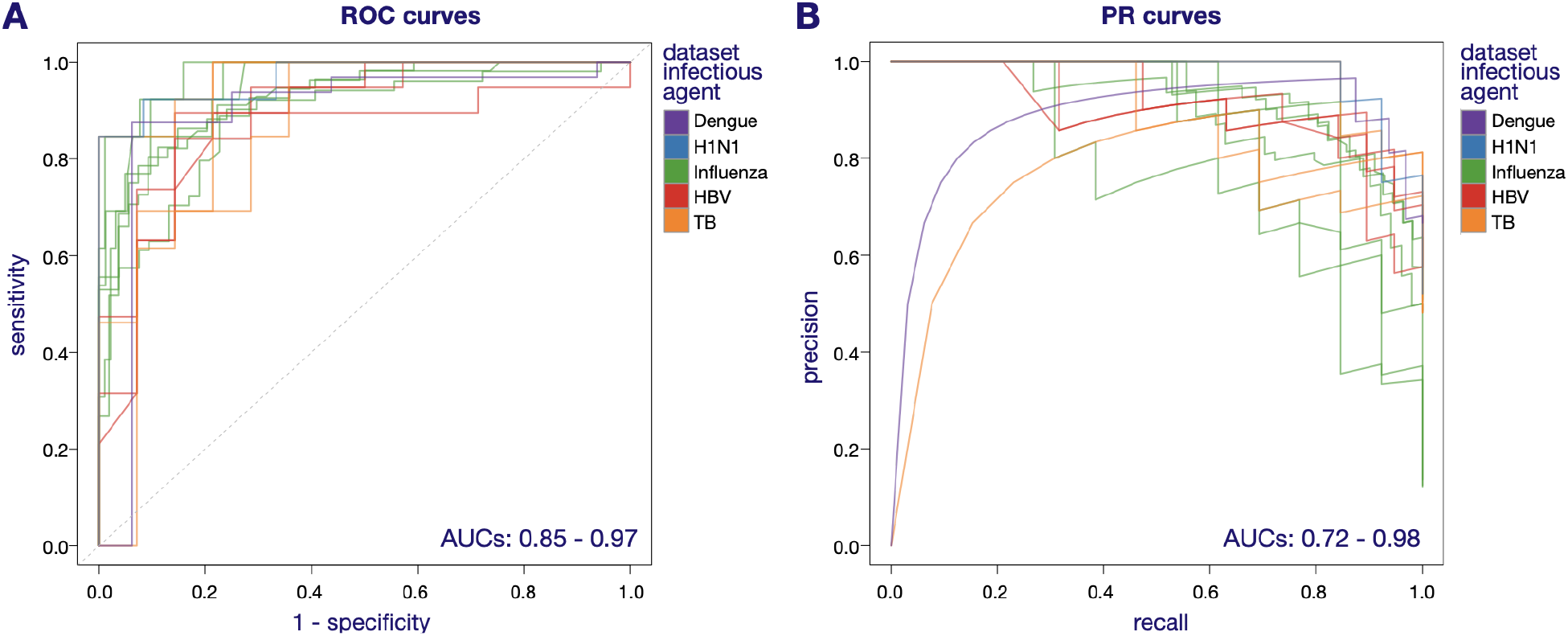
Optimized performance of transfer signatures. The classifying performance of the predicted phenotypes obtained from the random forest models using the transfer signature was assessed for each respective training dataset – where the transfer signatures were obtained from (**Methods, Suppl. Data S1**). **Panel A** displays the ROC curves. **Panel B** displays the PR curves. Each line depicts the curve obtained for a given training dataset. The lines are colored based on the infectious agent studied in the training dataset. ROC, Receiver operating characteristic. AUC, area under the curve. PR, precision recall.

By selecting genes for high importance from random forest models, transfer signatures become inevitably optimized and potentially overfit for each training study dataset. As we expect that fitting an overly expressive model will limit the generalizability of signatures to new datasets, it is important to note that, in downstream steps, transfer signatures consisted solely of a list of genes with no weights attached. This approach was undertaken so that error from over-fitting were not carried forward into the next level of analysis. Thus, in the next step we implemented dimension reduction by restricting the dataset to the genes of the transfer signatures and applying UMAP (Uniform Manifold Approximation and Projection) into two dimensions (but carrying over no details of the predictive model which selected the signature), followed by unsupervised clustering and a decision boundary to explore the generalization ability of transfer signature-based prediction on a new test dataset. Here below we present two use cases of this approach.

### Predictive power of transfer signatures in unseen data

The use of a transfer signature in an unsupervised approach, i.e., without retraining with known labels, is the most stringent implementation of the transfer signature, in particular when the training dataset is chosen to be only partially related to the target test dataset. In a first use case, we tested the hypothesis that information from a TB vaccine study in rhesus macaques can inform on progression from latent to active TB in a human cohort; i.e., inter-species application of a transfer signature. In the second use case, we tested whether information from severe dengue infection serves for the classification of cases of severe SARS-CoV-2 and of influenza A infection; i.e., inter-infectious disease transfer signature application (**Suppl. Data S1, Suppl. Fig. S1**). In these scenarios, the outcome is expressed as “enrichment”, i.e., the ability of a signature to increase the number of true cases in a population and “recall”, i.e., the fraction of cases that are retrieved in a population.

### Progression of latent to active tuberculosis

We modeled the value of the transfer signature obtained in an animal study to assess the challenge of enriching a clinical trial with individuals that are likely to reach a given endpoint. The scenario is the use of a pharmacological or vaccine intervention to prevent progression from latent tuberculosis to active disease. Progression to active tuberculosis is a rare event (estimated as 0.084 cases per 100 person-years) (*35*); therefore, it would be important to be able to recruit individuals that are the most likely to develop active infection within one year. Indeed, in the presence of a limited numbers of individuals that may reach a study endpoint the study may lack power to detect differences between the placebo and vaccine or treatment group.

Here, we tested transfer signatures obtained with the three time course datasets from Hansen et al. (*27*) (**Suppl. Data S1** – study 6). Effectively, this implies training of all literature signatures (excluding the cognate signature) on each of the three datasets, selecting the best performing genes for each respective dataset (**Fig. 1**). This study assessed the efficacy of a TB vaccine on rhesus macaques, with longitudinal samples from 27 rhesus macaques collected pre-vaccine, after vaccination but before TB challenge and four weeks post challenge. The phenotype used for training the random forest models was protection from TB (vaccine efficacy), defined as a computed tomography score of < 10 (protected, N=13) at any time point post challenge versus not protected (N=14).

We used as target dataset the data from Zak et al. (*36*), a longitudinal study assessing progression from latent to active TB (**Suppl. Data S1** – study 9). We defined cases as individuals that developed TB within a year (N=30) and controls as individuals that did not develop TB within a year after entry in the study (N=109; **Suppl. Data S1**). The results of the unsupervised clustering are shown in **Fig. 4**. With the transfer signature defined on the pre-vaccine rhesus macaque samples, 32.8% (22/67) of the predicted cases were true cases, i.e., developed active TB within a year, while the samples outside of this cluster contained only 11.1% (8/72) of true cases. Here, the unsupervised clustering led to a 3.0-fold enrichment (when comparing cases versus non-case cluster, or 1.5-fold enrichment when comparing the case cluster versus the general population) and a 73.3% recall. In a similar setting, but with the transfer signature derived from pre-challenge samples, we obtained a 2.0-fold enrichment (34.7% [17/49] versus 14.4% [13/90] when comparing cases versus non-case cluster, or 1.6 fold enrichment when comparing the case cluster versus the general population) and a 56.7% recall. With the signature derived from post-challenge samples, we obtained a 5.5-fold enrichment (60.0% [18/30] versus 11.0% [12/109] when comparing cases versus non-case cluster, or 2.8-fold enrichment when comparing the case cluster versus the general population) and 60.0% recall. Overall, the use of the transfer signature from an animal model would allow the prospective recruitment of individuals into clinical trials with a greater likelihood of reaching adequate study power.

**Fig. 4:**
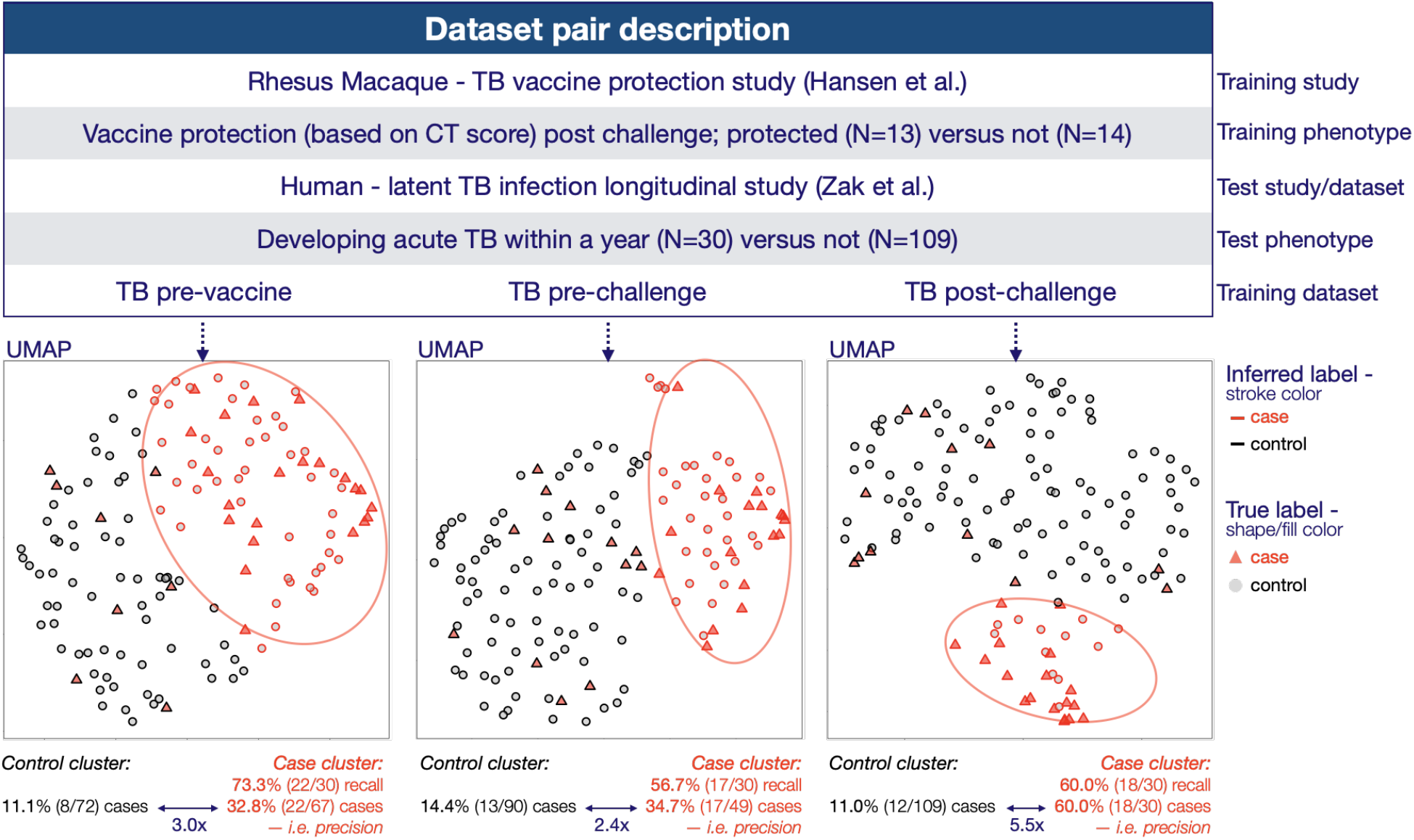
Tuberculosis progression use case – unsupervised clustering. **Top panel** displays the study design. **Bottom panel** displays the UMAP projection of the test dataset using the 50-gene-long transfer signature obtained from the respective training dataset shown in the top panel: pre-vaccine, pre-infectious challenge and post-challenge. Each sample is represented by a circle or a triangle. The stroke color indicates the inferred label (from the unsupervised clustering) and the shape and fill color indicate the true label. The recall and percentage of true cases in the different clusters (i.e. precision) is displayed below each UMAP projection. The colored circles surrounding the inferred case cluster are used solely for visual guidance. UMAP, Uniform Manifold Approximation and Projection. TB, Tuberculosis. CT, Computed Tomography scan.

### Severity of viral infection

We next assessed whether transfer signatures could be used in the setting of viral infection to predict or classify the severity of the symptoms of individuals that are hospitalized. Here, we tested transfer signatures obtained from the dataset from Devignot et al. (*23*) (**Suppl. Data S1** – study 1, children with acute dengue infection, with blood samples collected within 3 to 7 days after onset of fever). For the purpose of this analysis, we considered as cases the children with severe manifestations of disease (shock syndrome and hemorrhagic fever; N=32), while children that had uncomplicated dengue fever were considered controls (N=16). We then used data from Liao et al. (*37*) (**Suppl. Data S1** – study 7) and Dunning et al. (*8*) (**Suppl. Data S1** – study 8), as two different target datasets. The phenotypes were established at the time or before the RNA samples were obtained; therefore, the unsupervised clustering results (**Fig. 5)** reflect here the performance of transfer signatures as classifiers rather than predictors.

**Fig. 5:**
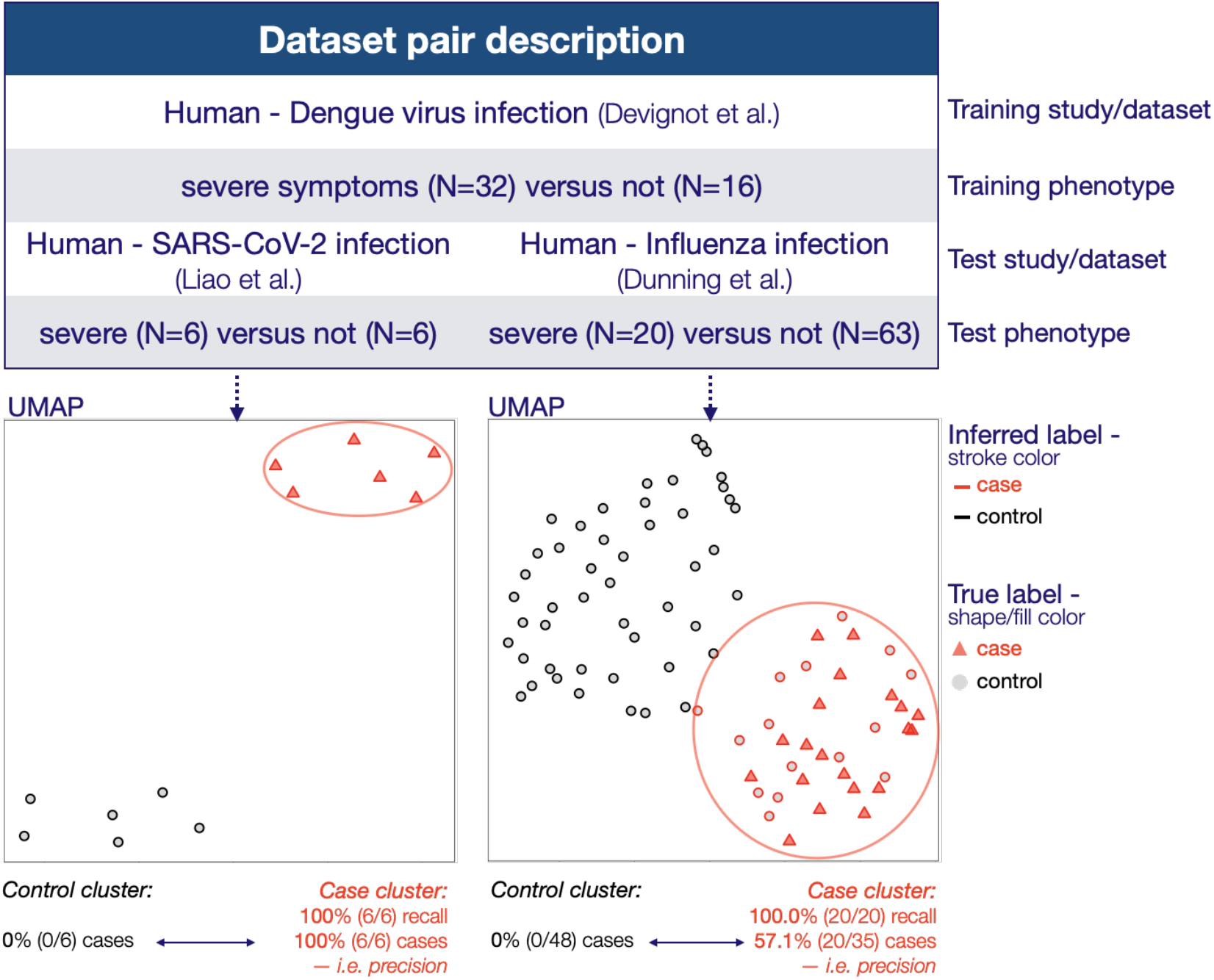
Severe viral disease use cases – unsupervised clustering. **Top panel** displays the study design. **Bottom panel** displays the UMAP projection of the test datasets using the 50-gene-long transfer signature obtained from the training dataset shown in the top panel. Each sample is represented by circle or triangle. The stroke color indicates the inferred label (from the unsupervised clustering) and the shape and fill color indicate the true label. The recall and percentage of true cases in the different clusters (i.e. precision) is displayed below each UMAP projection. The colored circles surrounding the inferred case cluster are used solely for visual guidance. UMAP, Uniform Manifold Approximation and Projection.

The study of Liao et al. (*37*) characterized bronchoalveolar lavage fluid immune cells from patients infected with SARS-CoV-2. For the purpose of this analysis, we considered as cases the individuals that were described in the original report as having severe disease (N=6), while individuals with moderate disease (N=3) or not infected (N=3) were considered as controls (total N=6). The RNA samples were obtained 4-10 days after the phenotypes were established. All true cases of severe SARS-CoV-2 study were correctly classified in unsupervised clustering; 100% precision and 100% recall and 2-fold enrichment when comparing the case cluster versus the general population (**Fig. 5**, left lower panel).

The study of Dunning et al. (*8*) characterized blood samples from individuals hospitalized with influenza. For the purpose of this analysis, we considered as cases the individuals that were described in the original report as requiring mechanical ventilation (N=20), while individuals that did not require mechanical ventilation were considered as controls (N=63). The inferred case cluster included 57.1% true cases (individuals that required mechanical ventilation), while none of the samples in the inferred control cluster were true cases, indicating a 2.4 fold enrichment when comparing the case cluster versus the general population and a 100% recall (**Fig. 5**, right lower panel).

Of note, the results displayed for the various use cases used a 50-gene-long transfer signature; however, similar results were obtained when selecting only the top 20 genes, while the performance dropped with some of the 10-gene transfer signatures (**Suppl. Fig. S6 and S7**). We obtained similar results when using transfer signatures derived with only hallmark signatures (**Data S1**) compared to transfer signatures based on all literature signatures (**Suppl. Fig. S8 and S9)**. Overall, both the SARS-CoV-2 and the influenza studies support the transfer of signatures across different viral infections to classify disease severity.

## Discussion

The present work considers a transcriptomic response as a complex set of expression profiles that includes genes uniquely modulated by a given perturbation, as well as shared responses (e.g. upregulation of interferon stimulated genes), cell type composition, prior history of infection and influence of genetics and the environment on the individual as well as experimental noise. A single study will inevitably sample from a complex transcriptome response to generate a study signature. Because many studies are limited in size, the resulting signatures will be, to some extent, the result of random sampling of a large space of deregulated genes and noise. Against this backdrop, a transfer signature aims at capturing informative markers of the broadest use.

Consistent with the working hypotheses, we found that many signatures in the literature are informative when tested in other datasets, including for related studies (e.g., same pathogen), or even across pathogens, vaccines and phenotypes. It is possible to make a parallel with transfer learning in the field of artificial intelligence where a model trained solving one task is repurposed on a related task. Transfer learning also uses information from larger compendia of datasets to inform and constrain the models. Our work and that of others also support the possibility of extending prediction across species; for example, a recent study indicates that the mouse transcriptome reveals potential signatures of protection and pathogenesis in human tuberculosis (*38*). The approach also found significant information content in more general sets of signatures, such as those listed in the hallmark gene sets of the Molecular Signatures Database, a result consistent with recent observations (*39*) and the use of co-expression patterns to define transcriptome modules (*40*). We interpret this as indication that even in the absence of pre-existing information in the literature, well-defined sets of hallmark genes will enable the extraction and creation of transfer signatures. Similarly, we observed that signatures of cell type composition (*22*) have also classifying power and could possibly inform on mechanisms of pathogenesis.

It is possible that the collections of signatures could serve as basis to explore human immunophenotypes: the baseline conditions that associate with a diversity of responses in human when exposed to, for example, infection or vaccines (*41*). The definition of human immunophenotypes currently requires complex genetic and immunological phenotyping (*20, 21*). In the use cases, a number of the transfer signatures were trained in samples at “baseline” – for example, prior to vaccination, at hospital admission, or before the development of disease – and still generated good prediction of distant endpoints. Thus, our work defines a general approach that creates transfer signatures to support immunophenotyping. A related concept is the interesting scenario of vaccine repurposing that has been recently discussed in the context of vaccines for SARS-CoV-2 infection (*42*). This concept implies that the general or broad responses to a given stimulus, here an unrelated antigen, may confer protection to a second pathogen by means of common or shared responses.

In future direction, there is the possibility to apply this concept at scale to determine when transfer works; i.e., what type of similarity must be present when transferring information, what are the determinants of successful transfer signatures. An important implication of this approach is that knowledge of the pathogenesis of diverse diseases may then be applicable, assuming transfer signature are predictive, to different diseases. This might have the effect of leveraging the depth of understanding of some diseases to explore less well understood diseases. A final goal is to define how many discrete and distinct transfer signatures/immunophenotypes can be defined and whether there are combinations of such elemental immunophenotypes. In summary, using machine learning approaches, we established the feasibility to transferring optimized gene shortlists from multiple studies to a target study with retention of predictive and classifier power. This could significantly facilitate the use of signatures for prospective studies before disease- and case-specific signatures can be determined.

## Data Availability

Data and code will be made available in Github upon publication

## Acknowledgments

We thank C. Maher and X. Ding for commentaries;

## Author contributions

J.di I. and A.T. conceived the project, J. di I. built the analytical models, I.B. and R.S. contributed analyses and pipelines, H.W.V. and A.T. provided biological grounding to the work. A.T. wrote the manuscript with support of all coauthors;

## Data and materials availability

Data and code availability will be made available in GitHub upon publication.

## Supplementary Materials

## Materials and Methods

### Gene signatures

We used five categories of signatures from publications, referred as “*literature signatures*”: (i) curated sets of gene lists –referred as *hallmark signatures* (N=50, https://www.gsea-msigdb.org/gsea/msigdb/collections.jsp) (*1*), (ii) gene signatures associated with cell composition in PBMC – referred as *cell type signatures* (N=22) (*2*), (iii) vaccine protection and response signatures –referred as *vaccine signatures* (N=13), (iv) progression from latent to active TB infection signatures –referred as *TB signatures* (N=20) and (v) viral and bacterial infection signatures –referred as *infection signatures* (N=43). All signature descriptions, sources, references and gene lists are provided in **Suppl. Data S1** and compiled in part in ref (*3*). Of note, due to gene nomenclature conversion issues, some signatures may be missing some genes identified in the parent paper.

### Training datasets

We used 14 different training datasets from six studies: one study on dengue infection (*4*) (**Suppl. Data S1** – study 1), one study on influenza H1N1 infection (*5*) (**Suppl. Data S1** – study 2), one study on trivalent Influenza vaccination comprising two cohorts, one with males (**Suppl. Data S1** – study 3) and one with females (*6*) (**Suppl. Data S1** – study 4) – each comprising 3 datasets obtained at different timepoints (pre-vaccination, day 1 and day 14 post-vaccination), one study on hepatitis B virus (HBV) vaccination (*7*) (**Suppl. Data S1** – study 5) – comprising 3 datasets obtained at different timepoints (pre-vaccination, day 3 and day 7 post-vaccination) and one study on tuberculosis (TB) vaccination in rhesus macaques (*8*) (**Suppl. Data S1** – study 6) – comprising 3 datasets obtained at different timepoints (pre-vaccination, pre-challenge with TB and 28 days post-challenge with TB). Of note, several studies contain multiple non-independent datasets (or timepoints). This design is expected to help understand the biology of transfer transcriptome signatures and to monitor what are the earliest time points with predictive power.

### Test datasets

We used 3 test datasets from three studies: one study on bronchoalveolar lavage in SARS-CoV-2 infection (*9*) (**Suppl. Data S1** – study 7), one study on influenza infection (*10*) (**Suppl. Data S1** – study 8) and one longitudinal study on TB progression in latently infected individuals (*11*) (**Suppl. Data S1** – study 9). Of note, all test datasets were independent from each other and from any training datasets.

### Phenotypes used

We explored multiple phenotypes in the training and test datasets, that can be categorized in four groups, namely (i) severity of symptoms during viral infection (for dengue, influenza and SARS-CoV-2 infection studies), (ii) vaccine response (for both HBV and influenza vaccination studies), (iii) disease state - for TB vaccination study in rhesus macaque, and (iv) time to disease in the longitudinal study TB progression. Further description and the number of individuals in each phenotype category per study are provided in **Suppl. Data S1**. Of note, the phenotype extracted from the publicly available datasets is not necessarily the one used in the original study. The differences in phenotype definition, if any, are provided in **Suppl. Data S1**. As an example, we used categorical/binary phenotypes even when the original study used numerical phenotype in order to be consistent across datasets and to better mimic future potential practical use cases.

### Gene Signature evaluation in training datasets

A random forest model was run on each “literature signature – training dataset” pair (hereafter referred as S-D pair). In order to prevent overfitting the model to a specific pair and given the downstream goal of identifying genes that were common biomarkers across experiments and conditions, rather than specific to a single study or pair, hyperparameters were not tuned and were used as follow: number of trees (N=1,000); all other hyperparameters were the default in *randomForest* function from the R package “randomForest” (https://cran.r-project.org/web/packages/randomForest/index.html). In the model, normalized gene expression of the subset of genes present in the signature was used to classify the phenotype of interest. For RNAseq input datasets, the normalization consisted in log10 (reads per million mapped read + 1e-7) and genes with less than 20 reads in every sample in the dataset were removed. For microarray input datasets, we retrieved the normalized data from the GEO repository, averaged the normalized signal of all probes per gene and finally used the log10 (average normalized signal per gene + 1e-7) as input for the model. The code used for running the random forest modeling was adapted from https://github.com/jasonzhao0307/R_lib_jason/blob/master/RF_output.R

Given the small sample size of most datasets, the models were trained using leave-one-out cross validation (LOOCV), where for each sample of a dataset, all other samples from the same dataset are used to train the RF model, and the resulting model is used to predict the label or phenotype of the remaining sample. The LOOCV strategy results in one RF model trained per sample per S-D pair. To obtain the combined gene importance feature for a specific S-D pair, the gene importance scores were averaged across all models from a given S-D pair, resulting in one score of “importance” per gene per S-D pair, where the importance measure reflects the mean decrease in node impurity. The receiving operating characteristic (ROC) and precision recall (PR) area under the curve (AUC) are computed using the scores of the single left-out sample per trained model.

### Extraction of transfer signatures

Only literature signatures that had a ROC AUC percentile above a given threshold were used at this step. Percentiles were determined as follows: for each S-D pair, 100 random gene lists of the same size were used to compare the performance of the literature signature. Percentiles were used to be able to compare the numbers across datasets that did not have the same case/control distributions. The thresholds of 70, 80 and 90 were empirically tested (**Suppl. Fig. S10**) and the 70^th^ percentile was chosen, as the two latter were too stringent (in terms of number of literature signatures that passed the threshold) when the signatures were split by group. In order to be able to compare the gene importance feature across literature signatures for a given training dataset, each gene literature signature importance feature was standardized to obtain a mean of 0 and a standard deviation of 1 (z-scores). The z-scores were then aggregated, and the top unique genes were selected as representing the transfer signature.

The number of genes in a transfer signature (N=10, 20 and 50) were empirically tested (**Suppl. Fig. S3**). The size of 50 genes was chosen for further analyses, with the rationale that (i) 50 genes appeared to provide the best performance in the datasets for which the signature length appeared to play the largest impact and (ii) the larger the signature length the more likely the signature will generalize to other datasets under different conditions. We did however not test transfer signatures containing more than 50 genes for practicality purposes, as the foreseen use of transfer signature will not necessarily be associated with high-throughput sequencing, and a limited size signature has the potential to be more broadly applicable (for example if the markers are assessed through qPCR rather than RNAseq).

Throughout the main text, we used the transfer signature derived from all contributing literature signatures (**Fig. 3**-**5**), but we also generated and tested transfer signatures based on hallmark and cell type signatures to assess whether they could be broadly applicable, see **Suppl. Fig S4-9**. The gene lists of transfer signatures are provided in **Suppl. Data S1**.

### Prediction in unseen test dataset

Genes identified as markers of “commonality” – present in transfer signatures - were used in an unsupervised analysis to cluster samples from new test datasets, that originated from independent studies (notably new condition, new organism or new infectious agent). The dimension reduction was performed using Uniform Manifold Approximation and Projection (UMAP), followed by Hierarchical Density-Based Spatial Clustering of Application with Noise (HDBSCAN) (*12*) which can cluster data of varying shape and density. In this approach, the only parameter required is the minimal number of samples per cluster. For this purpose, we tested empirically the minimal number, by identifying the number of samples per cluster that resulted in the lowest number of outliers multiplied by a penalty score equivalent to the square of the number of clusters. This approach limits the creation of excessive numbers of clusters, which could make interpretation difficult. The minimum number of samples per cluster was set to contain at least 7% of the total population. HDBSCAN was run using the *hdbscan* command from the R package “dbscan” (https://github.com/mhahsler/dbscan). The samples considered as outliers by HDBSCAN, were attributed to the closest cluster label using the 3 nearest neighbors with the *knn* command from the R package “dbscan” (https://github.com/mhahsler/dbscan). The code used for running the dimensionality reduction and unsupervised clustering was adapted from https://github.com/NikolayOskolkov/ClusteringHighDimensions/blob/master/easy_scrnaseq_tsne_cluster.R

Once the clusters were identified, the inference of cluster attribution (case or control) was estimated based on the expression of the genes in the signature. Specifically, we did not directly compare the expression between training and test set as the range of expression is most likely more different across dataset than across phenotype within a dataset. We used the direction of the signal rather than the absolute value: for each gene present in the transfer signature, we compared the median expression in each cluster and recorded the direction of the signal in each cluster (high, low or intermediate - in the presence of more than 2 clusters). We performed the same in the training dataset where the transfer signature was obtained from, using the true labels (case/control) instead of clusters to group the samples. We then assessed which cluster in the test dataset had the highest proportion of genes that matched the label of interest in the training dataset (in terms of signal direction) and defined it as “case cluster”, while the other cluster(s) were defined as control cluster. In the rare case where two clusters had the same proportion of matches, we compared the sum of the absolute difference (in median expression) of the genes that matched the direction of the signal in the training dataset. Of note, we need to use biological understanding to decide which phenotype label in the training dataset (**Data S1**) would resemble the most the phenotype of interest (“case”) in the test dataset, if not the clusters might be inverted. For example, in the tuberculosis use case, when we used the transfer signatures obtained with the post-challenge timepoint, we expected that the rhesus macaques that were not protected by the vaccine at the end of the study, were the most likely to resemble the individuals that were going to develop acute TB within in a year, as the rhesus macaques were already in a disease state at that time point and the unprotected animals were expected to have a much higher level of immune gene expression in the disease state. While on the opposite, when we used the transfer signatures obtained from the pre-vaccine or pre-challenge datasets, we expected the “case” phenotype to the be rhesus macaques that were protected by the vaccine at the end of the study, as the animals with higher basal level of immune gene expression (such as interferon stimulated genes) are expected to have a higher likelihood of vaccine protection.

---

**Suppl. Fig. S1.**
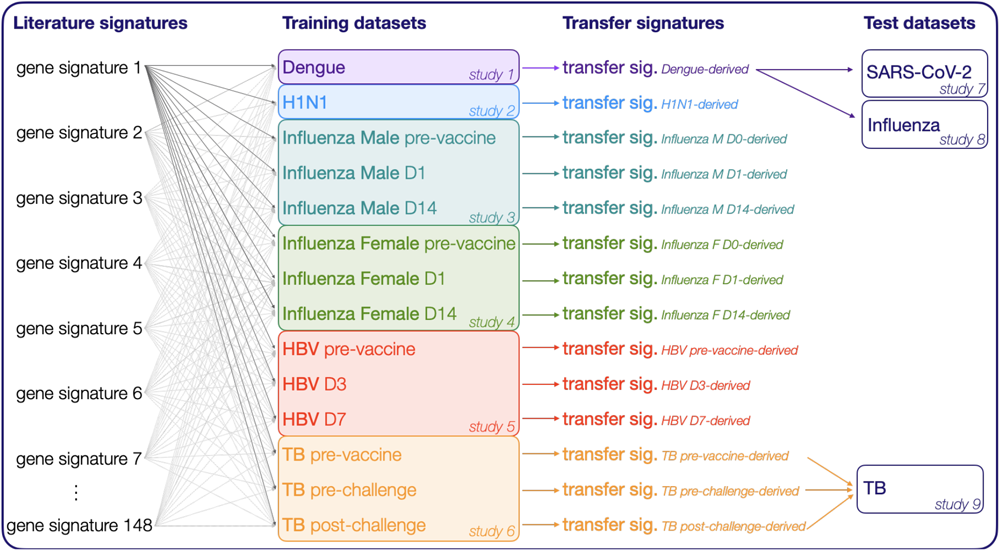
Study design – pairing of signatures, training and test datasets. This figure complements the study design depicted in **Fig. 1**. The schema highlights the pairing of signatures and datasets used for training to generate the transfer signatures and finally the pairing on transfer signatures and test datasets. From left to right: each literature signature (N=148) is used with each training dataset (N=14) as an input to train a random forest model (see **Fig. 1A**). In other words, there are 148 random forest models per training dataset. The gene importance feature and ROC AUC from all random forest models obtained for a given training dataset is used as input to generate one “*transfer signature*” per training dataset (**Fig. 1B**). In other words, a single transfer signature is obtained by combining the information obtained from a set of literature gene signatures (here, we start with all literature signatures, except the cognate signature – signature coming from the same paper than the dataset – for a given training dataset). Finally, the transfer signature derived from each training dataset can be used as an input for unsupervised clustering of a new test dataset (see **Fig. 1C**). The pairings between transfer signatures and test datasets used in this study are depicted by the arrows. Description of signatures and datasets are provided in **Suppl. Data S1**. D0, Day 0 is equivalent to pre-vaccine. D1, Day 1. D3, Day 3. D7, Day 7. D14, Day 14. F, Female. M, Male.

**Suppl. Fig. S2.**
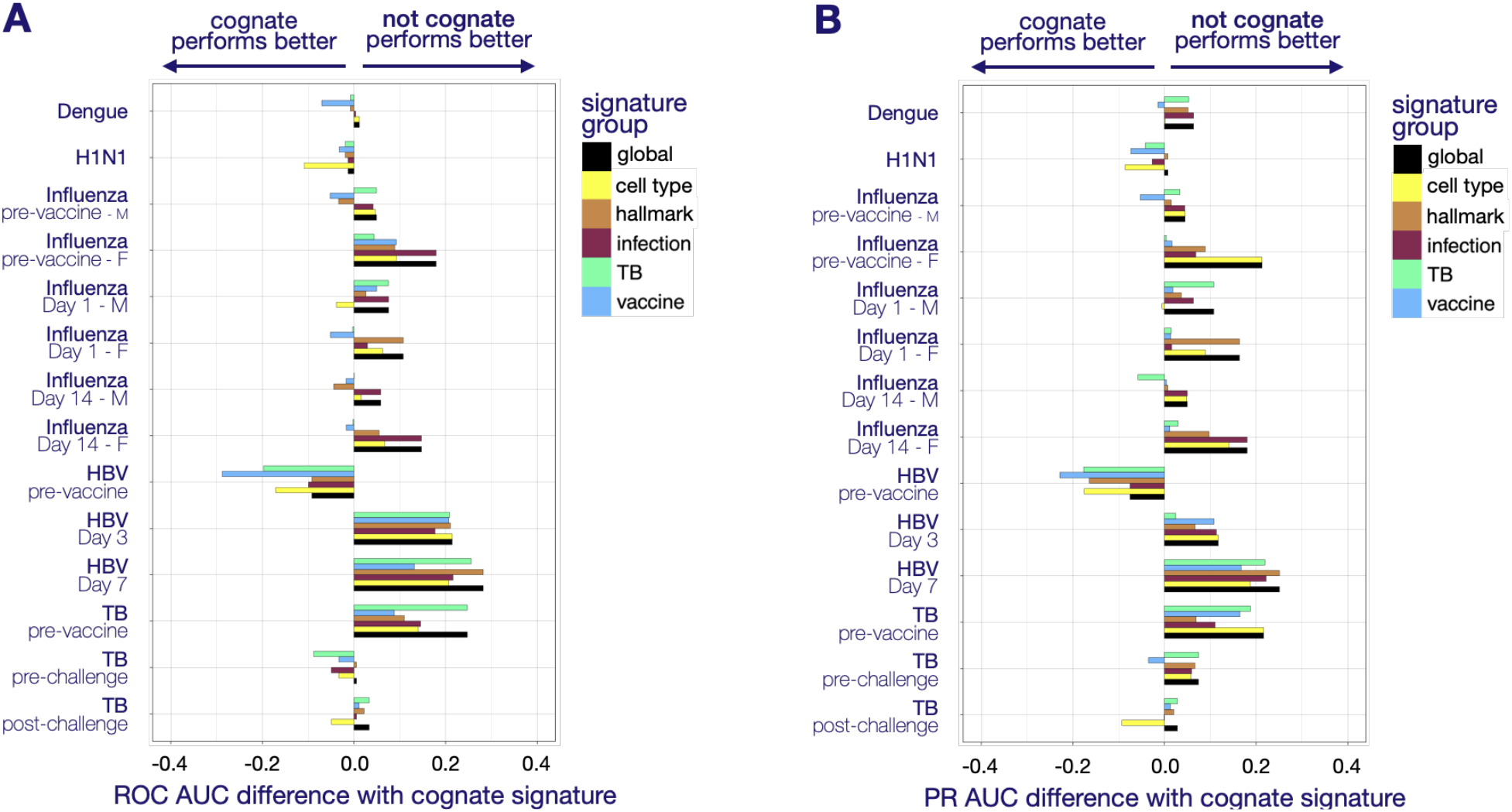
Performance of literature signatures as compared to cognate signatures. The classifying performance of the predicted phenotypes obtained from the random forest leave-one-out cross validation strategies using the literature signatures was assessed for each training dataset (**Methods, Suppl. Data S1**). Both panels display the difference in performance (as measured in ROC AUC – **Panel A** – or PR AUC – **Panel B**) between the cognate signature (signature from the same paper than the dataset) and the best performing signature from the literature. When there were multiple signatures originate from the same paper than the training dataset the best performing one was used as “cognate”. The literature signatures that outperformed the cognate signature have a positive difference and inversely the ones that did not perform as well have a negative difference. The results are depicted for each group of signatures (**Methods, Suppl. Data S1**) – ‘*global*’ encompasses all groups of signatures. The color code is provided in the legend. For both panels, there could be multiple reasons why the cognate signatures do not necessarily perform the best, including (i) the phenotype used for this study may differ from the parent study (f.ex. categorical phenotype rather than numerical; **Suppl. Data S1**), (ii) the cognate signature may only apply to one timepoint of the parent dataset, and finally (iii) some genes from the cognate signature may not have been retrievable due to gene nomenclature conversion issue (**Suppl. Data S1**). In any case, this validates that minor changes in the conditions or analytical settings can alter the results, supporting our strategy to focus on what is shared and transferable across studies rather than what is specific to a single study or condition. ROC, Receiver operating characteristic. AUC, area under the curve. PR, precision recall.

**Suppl. Fig. S3.**
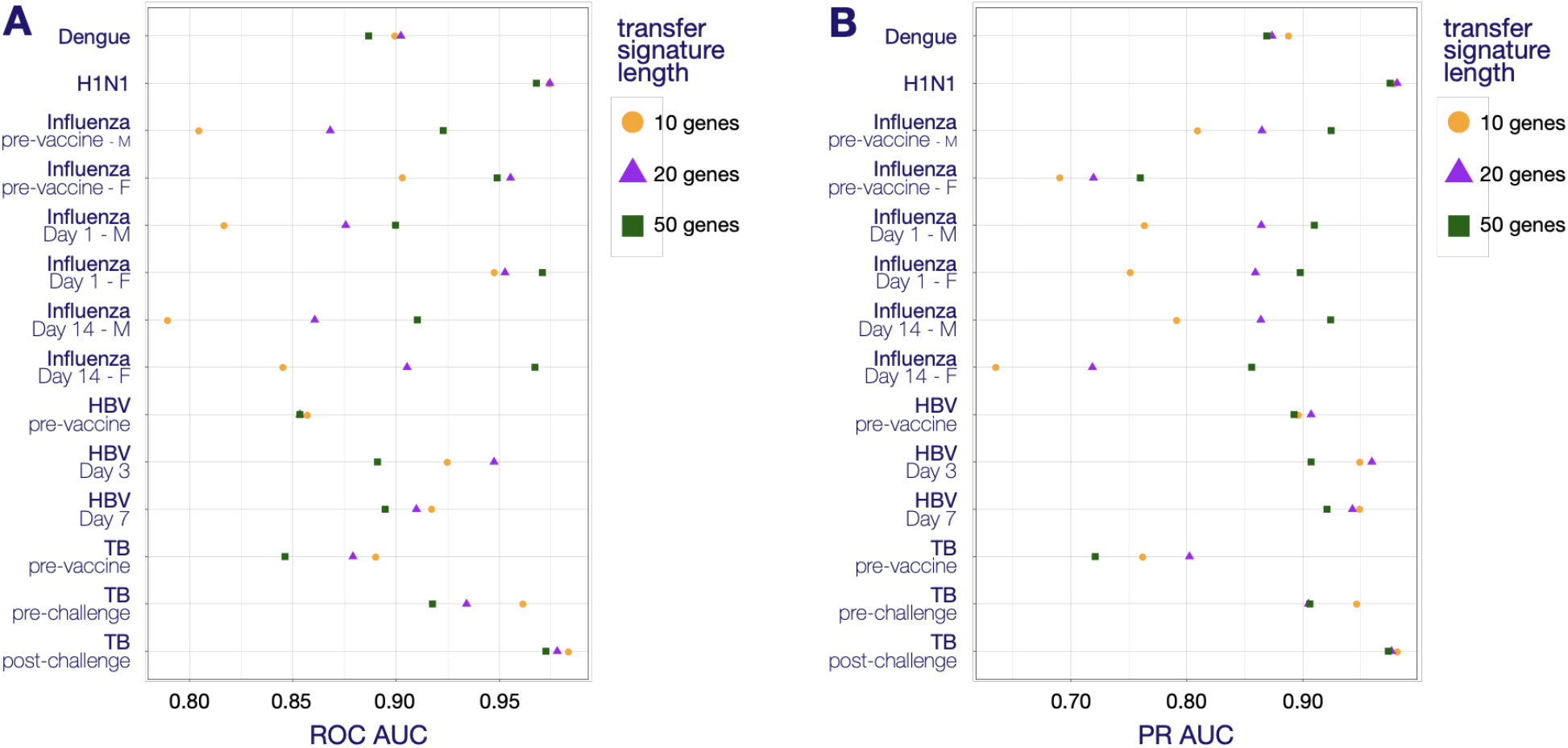
Performance of transfer signatures of different sizes. The classifying performance of the predicted phenotypes obtained from the random forest models (with leave-one-out cross validation) using transfer signatures of varying sizes was assessed for each respective training dataset – where the transfer signatures were obtained from (**Methods, Suppl. Data S1**). Three lengths of transfer signatures are depicted in different color and shape. The color code is provided in the legend. **Panel A** displays the ROC AUC obtained for each training dataset. **Panel B** displays the PR AUC obtained for each training dataset. The size of 50 genes was chosen for further analyses, with the rationale that (i) 50 genes appeared to provide the best performance in the datasets for which the transfer signature length appeared to play the largest impact and (ii) the larger the signature length the more likely the signature will generalize to other datasets with different conditions. We did not test transfer signatures containing more than 50 genes for practicality purposes, as the foreseen use of transfer signature will not necessarily be associated with high-throughput sequencing. ROC, Receiver operating characteristic. AUC, area under the curve. PR, precision recall.

**Suppl. Fig. S4.**
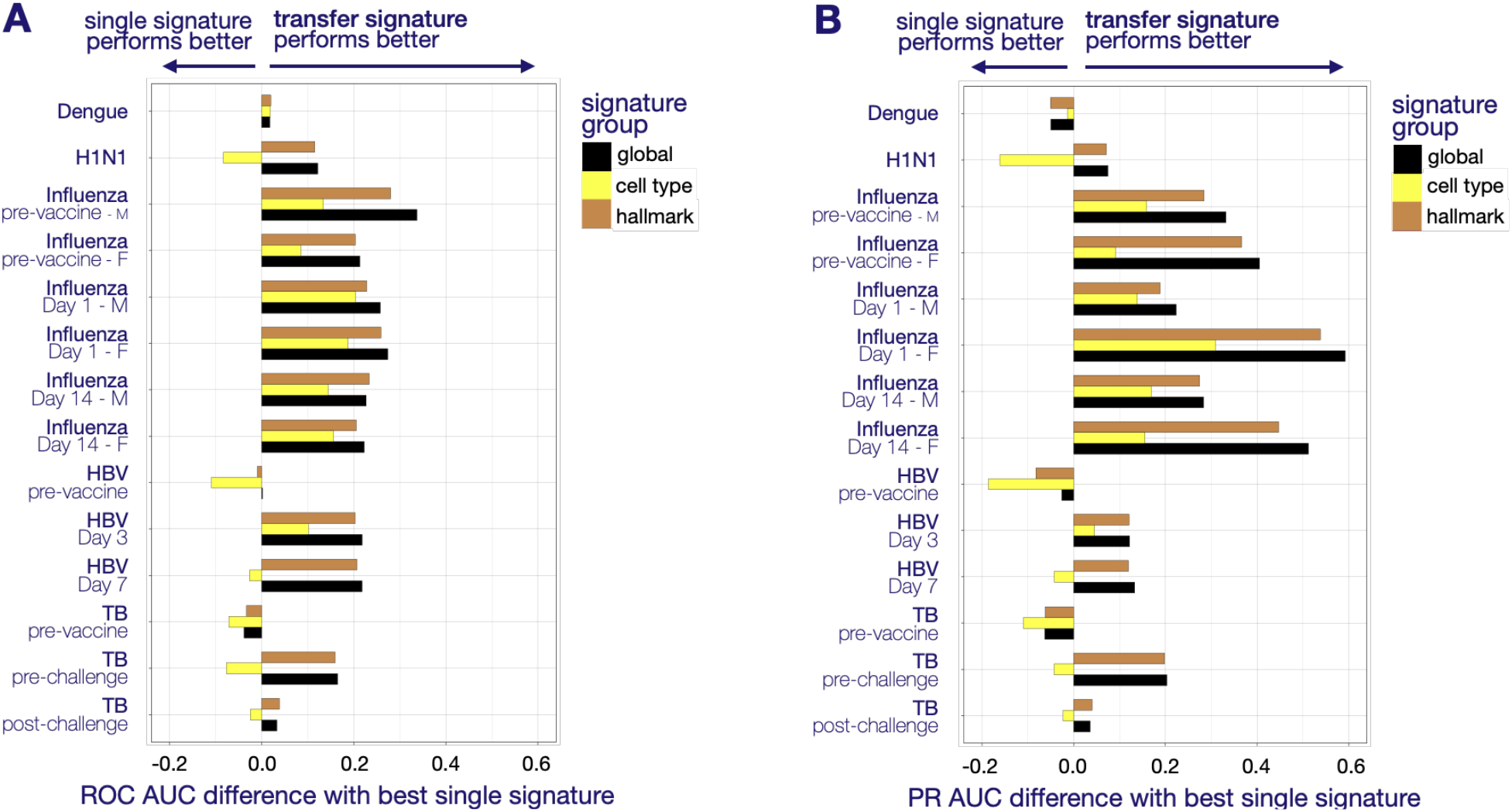
Performance of transfer signatures as compared to single signatures. The classifying performance of the predicted phenotypes obtained from the random forest models (with leave-one-out cross validation) using the transfer or single literature signatures was assessed for each training dataset (**Methods, Suppl. Data S1**). Both panels display the difference in performance (as measured in ROC AUC – **Panel A** – or PR AUC – **Panel B**) between the transfer signature and the best single performing literature signature (including the cognate signature for the dataset). The transfer signatures that outperformed the best single literature signature have a positive difference and inversely the ones that did not perform as well have a negative difference. For the purpose of this analysis, we developed not only one transfer signature per training dataset (that was obtained when starting with all literature signatures, **Suppl. Fig. S1**), but also one transfer signature for the *cell type* and *hallmark* group of signatures, per training dataset. In other words, we started with different subsets of literature signatures to compute the transfer signature and the results are depicted for those three groups of signatures (**Methods, Suppl. Data S1**) – ‘*global*’ encompasses all signatures. The color code is provided in the legend. In most instances, the transfer signature outperforms the best performing single signature, with the advantage of increasing the likelihood of generalization in new datasets as transfer signatures are obtained from multiple literature signatures, reducing the risk of extracting condition/study specific markers. ROC, Receiver operating characteristic. AUC, area under the curve. PR, precision recall.

**Suppl. Fig. S5.**
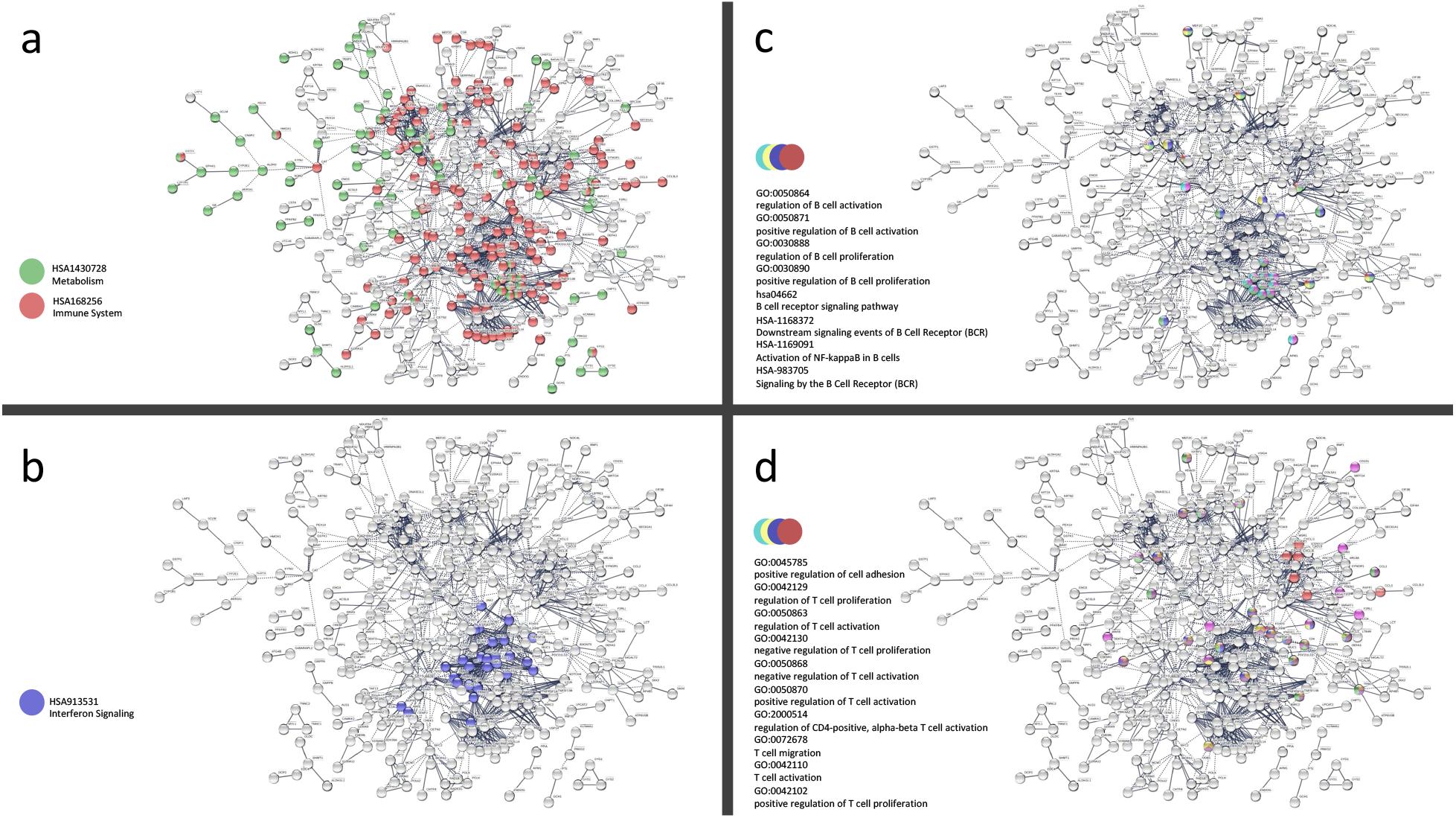
Functional enrichment for Transfer Signatures. Genes from the 14 transfer signatures (total unique genes: n=641, **Suppl. Data S1**) were assessed using STRING EMBL (https://string-db.org). The four panels present a network of high confidence interactions (pruning nodes/genes with no connections). The network has significantly more interactions than expected, PPI enrichment p-value < 1.0e^-16^. **Panel a** highlights the importance of genes of metabolism (green) and immunity (red) in transfer signatures. **Panels b-d** show that the transfer signatures effectively map the relevant parts of the immune system including innate (**b**, emphasis on interferon signaling) and adaptive responses (**c**,**d**, emphasis on T and B cell processes).

**Suppl. Fig. S6.**
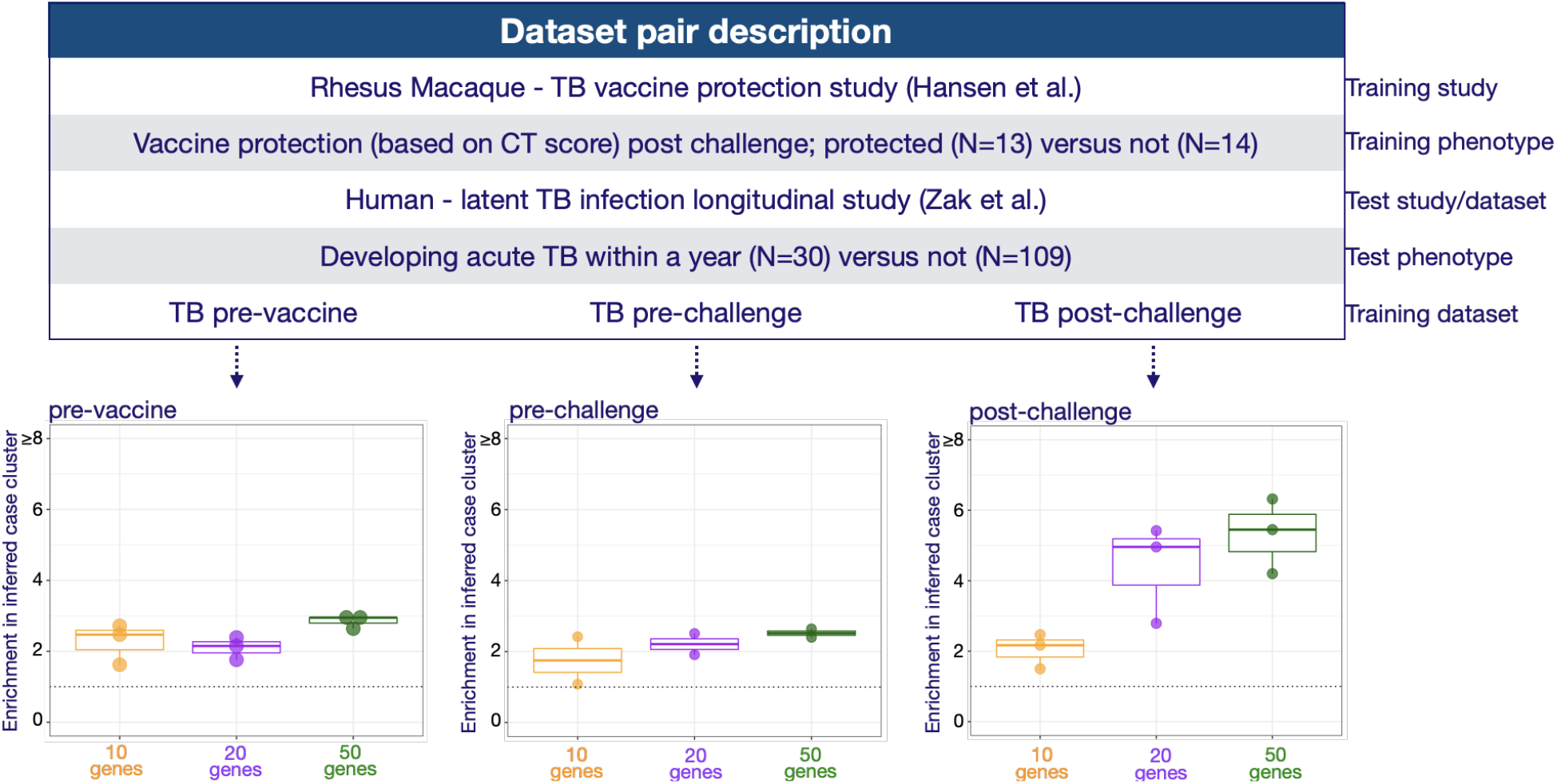
Tuberculosis progression use case – comparison of transfer signature size performance. **Top panel** shows the study design as displayed in **Fig. 4. Bottom panel** displays the enrichment of cases in the inferred case cluster compared to the other cluster(s) – y axis – using transfer signatures of differing size – x axis. The three plots represent the results obtained with transfer signatures trained with samples obtained at 3 different timepoints shown in the top panel: pre-vaccine, pre-infectious challenge and post-challenge. The results are depicted as boxplot with the individual data overlaid, where each dot represents the result obtained with a transfer signature derived from a different group of literature signatures (global, cell type and hallmark), as explained in **Suppl. Fig. S4** (see also **Methods, Suppl. Data S1**). The enrichment per transfer signature group is further detailed for the 50-gene-long transfer signatures in **Suppl. Fig. S8**.

**Suppl. Fig. S7.**
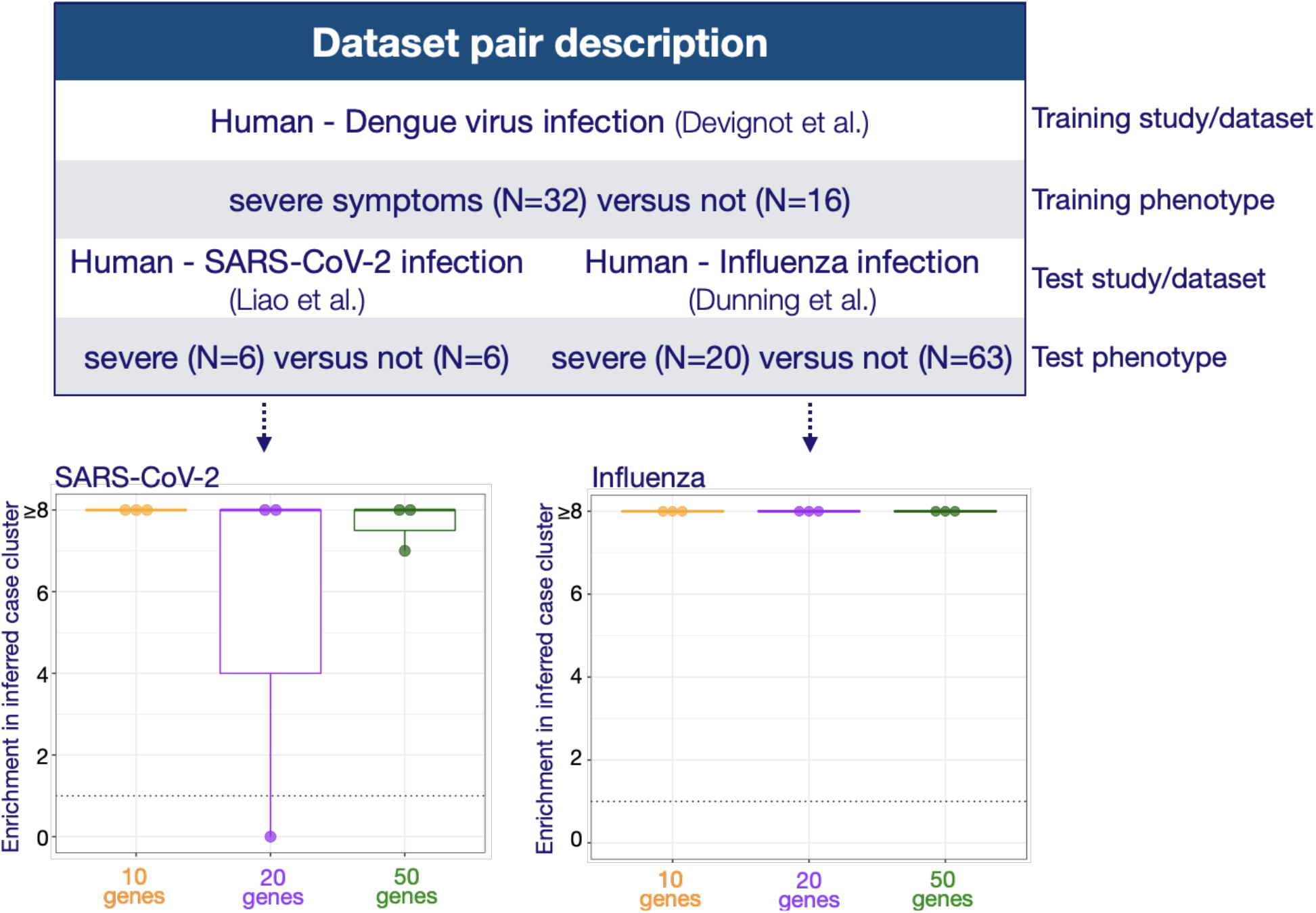
Severe viral disease use cases – comparison of transfer signature size and performance. **Top panel** shows the study design as displayed in **Fig. 5. Bottom panel** displays the enrichment of cases in the inferred case cluster compared to the other cluster(s) – y axis – using transfer signatures of differing size – x axis. The results are depicted as boxplot with the individual data overlaid, where each dot represents the result obtained with a transfer signature derived from a different group of literature signatures (global, cell type and hallmark), as explained in **Suppl. Fig. S4** (see also **Methods, Suppl. Data S1**). The enrichment per transfer signature group is further detailed for the 50-gene-long transfer signatures in **Suppl. Fig. S9**. Enrichment below 1 indicates that the “case” cluster was inversely inferred (**Methods**). Here, enrichment depicted as ≥8 indicate that all cases were correctly labeled/present in the inferred case cluster, as seen in **Fig. 5**.

**Suppl. Fig. S8.**
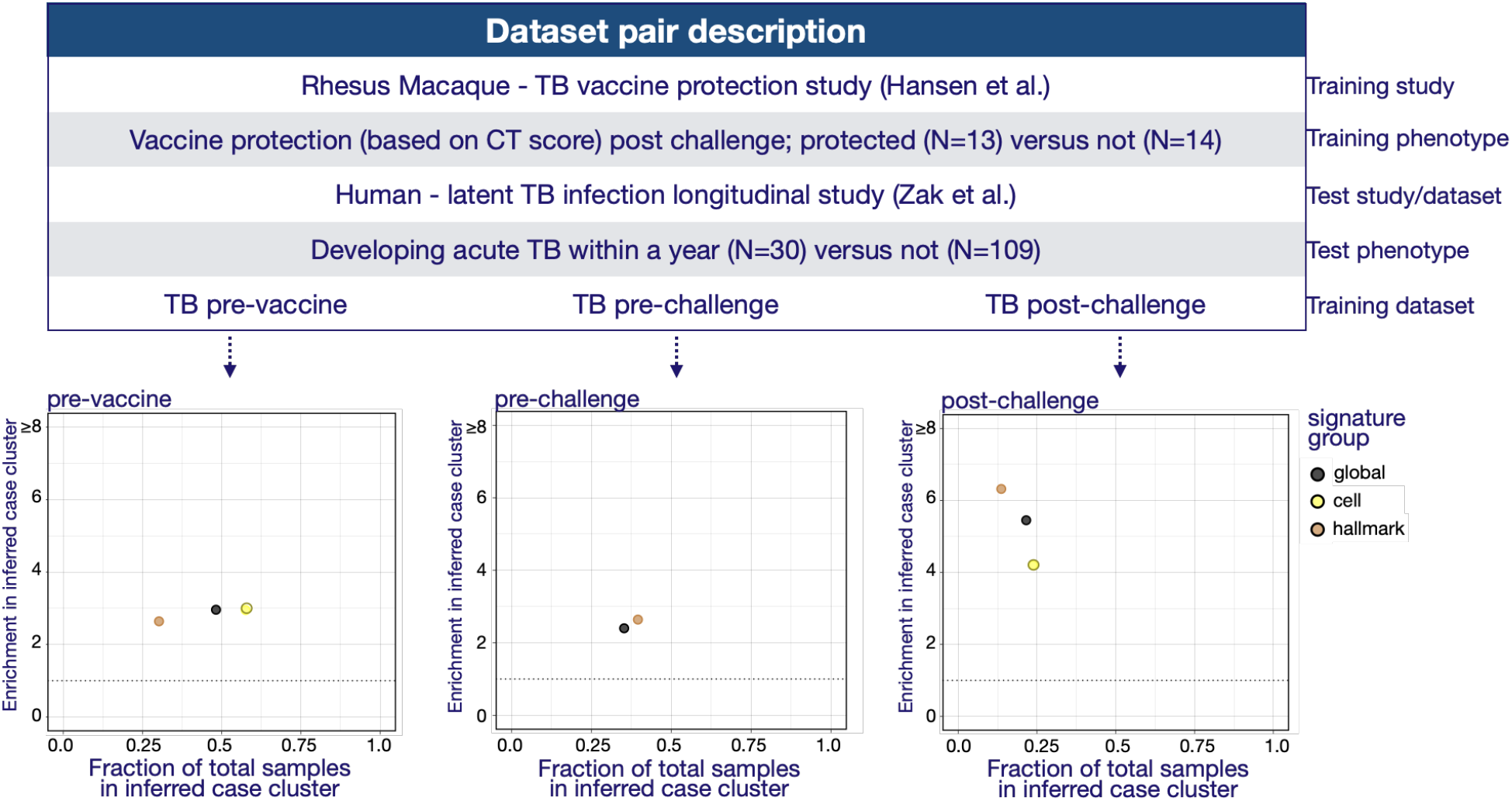
Tuberculosis progression use case – comparison of transfer signatures obtained from different signature groups. **Top panel** shows the study design as displayed in **Fig. 4. Bottom panel** displays the enrichment of cases in the inferred case cluster compared to the other cluster(s) using 50-gene-long transfer signatures – y axis – versus the fraction of samples present in the inferred case cluster – x axis. The three plots represent the results obtained with transfer signatures trained with samples obtained at 3 different timepoints shown in the top panel: pre-vaccine, pre-infectious challenge and post-challenge. Each dot represents the result obtained with a transfer signature derived from a different group of literature signatures (global, cell type and hallmark) – as explained in **Suppl. Fig. S4** and where ‘*global*’ encompasses all signatures (see also **Methods, Suppl. Data S1**). The color code is provided in the legend. The missing dot for the cell type transfer signature trained on the TB pre-challenge dataset indicates that there were not enough (<50) genes present in the signatures that passed the initial 70^th^ percentile threshold used to extract the transfer signature (**Methods**).

**Suppl. Fig. S9.**
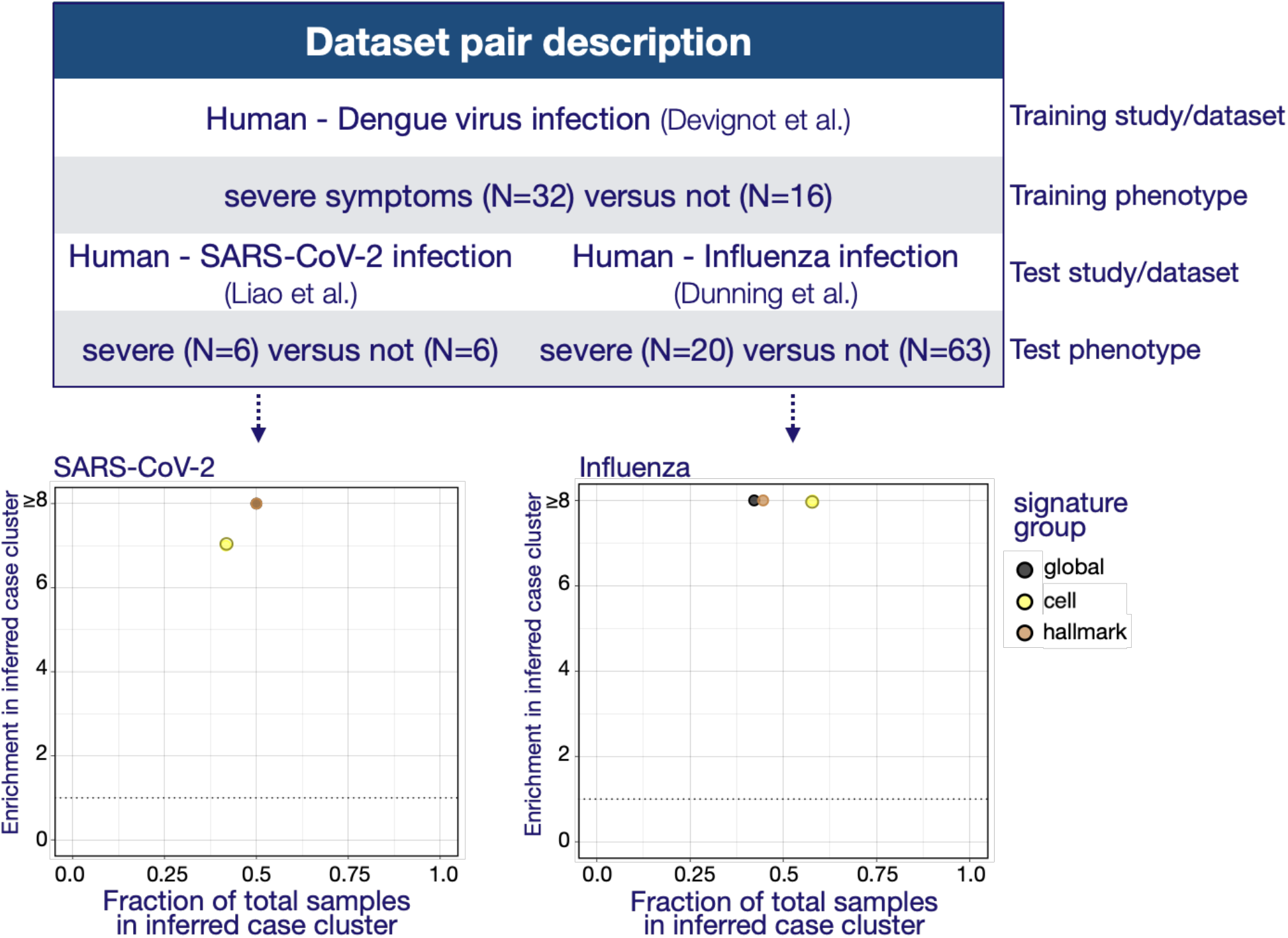
Severe viral disease use cases – comparison of transfer signatures obtained from different signature groups. **Top panel** displays the study design. **Bottom panel** displays the enrichment of cases in the inferred case cluster compared to the other cluster(s) using 50 gene commonality signatures – y axis – versus the fraction of samples present in the inferred case cluster – x axis. Each dot represents the result obtained with a transfer signature derived from a different group of literature signatures (global, cell type and hallmark) – as explained in **Suppl. Fig. S4** and where ‘*global*’ encompasses all signatures (see also **Methods, Suppl. Data S1**). The color code is provided in the legend. In the SARS-CoV-2 example, due to the small sample size, multiple transfer signatures obtained from different groups of signatures (global and hallmark) generated the same clustering, yielding to the same results in terms of enrichment and fraction and are therefore overlaid and non-visible individually. Here, enrichments depicted as ≥8 indicate that all cases were correctly labeled/present in the inferred case cluster, as seen in **Fig. 5**.

**Suppl. Fig. S10.**
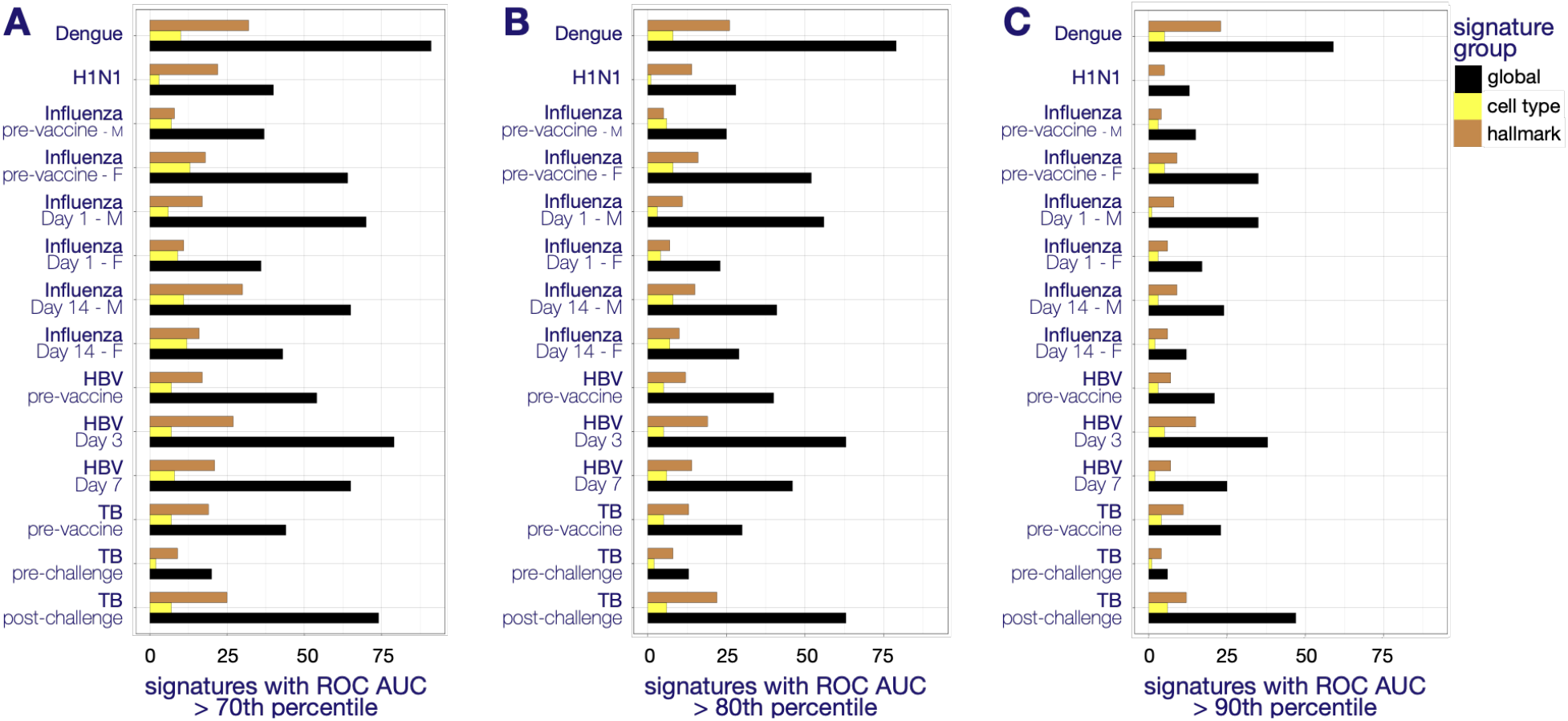
Number of literature signatures at different percentile threshold. The barplots display, for the three groups of signatures used to generate transfer signatures (global, cell type and hallmark), the number of signatures with ROC AUC higher than the 70^th^ percentile (**Panel A**), 80^th^ percentile (**Panel B**) and 90^th^ percentile (**Panel C**) for each signature group. The classifying performance of the predicted phenotypes are obtained from the random forest models (with leave-one-out cross validation) using the literature signatures was assessed for each training dataset. The percentiles are obtained by comparing the literature signature performance to 100 random gene lists of the same size. The higher the percentile, the better the performance of the signature. The color code is provided in the legend. ROC, Receiver operating characteristic. AUC, area under the curve.

